# Effects of previous infection, vaccination, and hybrid immunity against symptomatic Alpha, Beta, and Delta infections

**DOI:** 10.1101/2023.04.21.23288917

**Authors:** Heba N. Altarawneh, Hiam Chemaitelly, Houssein H. Ayoub, Patrick Tang, Mohammad R. Hasan, Hadi M. Yassine, Hebah A. Al-Khatib, Asmaa A. Al Thani, Peter Coyle, Zaina Al-Kanaani, Einas Al-Kuwari, Andrew Jeremijenko, Anvar Hassan Kaleeckal, Ali Nizar Latif, Riyazuddin Mohammad Shaik, Hanan F. Abdul-Rahim, Gheyath K. Nasrallah, Mohamed Ghaith Al-Kuwari, Adeel A. Butt, Hamad Eid Al-Romaihi, Mohamed H. Al-Thani, Abdullatif Al-Khal, Roberto Bertollini, Laith J. Abu-Raddad

**Author notes:** Correspondence: Dr. Heba Altarawneh, or Professor Laith J. Abu-Raddad,.

## Abstract

**Background:** Protection against SARS-CoV-2 symptomatic infection and severe COVID-19 of previous infection, mRNA two-dose vaccination, mRNA three-dose vaccination, and hybrid immunity of previous infection and vaccination were investigated in Qatar for the Alpha, Beta, and Delta variants.

**Methods:** Six national, matched, test-negative, case-control studies were conducted between January 18-December 18, 2021 on a sample of 239,120 PCR-positive tests and 6,103,365 PCR-negative tests.

**Results:** Effectiveness of previous infection against Alpha, Beta, and Delta reinfection was 89.5% (95% CI: 85.5-92.3%), 87.9% (95% CI: 85.4-89.9%), and 90.0% (95% CI: 86.7-92.5%), respectively. Effectiveness of two-dose BNT162b2 vaccination against Alpha, Beta, and Delta infection was 90.5% (95% CI, 83.9-94.4%), 80.5% (95% CI: 79.0-82.0%), and 58.1% (95% CI: 54.6-61.3%), respectively. Effectiveness of three-dose BNT162b2 vaccination against Delta infection was 91.7% (95% CI: 87.1-94.7%). Effectiveness of hybrid immunity of previous infection and two-dose BNT162b2 vaccination was 97.4% (95% CI: 95.4-98.5%) against Beta infection and 94.5% (95% CI: 92.8-95.8%) against Delta infection. Effectiveness of previous infection and three-dose BNT162b2 vaccination was 98.1% (95% CI: 85.7-99.7%) against Delta infection. All five forms of immunity had >90% protection against severe, critical, or fatal COVID-19 regardless of variant. Similar effectiveness estimates were observed for mRNA-1273.

**Conclusions:** All forms of natural and vaccine immunity prior to Omicron introduction provided strong protection against infection and severe COVID-19. Hybrid immunity conferred the strongest protection and its level was consistent with previous-infection immunity and vaccine immunity acting independently of each other.

## INTRODUCTION

Prior to introduction of severe acute respiratory syndrome coronavirus 2 (SARS-CoV-2) Omicron (B.1.1.529) variant in December of 2021 [1], Qatar experienced three waves dominated sequentially by the original virus [2], Alpha (B.1.1.7) variant [3], and Beta (B.1.351) variant (Supplementary Figure S1) [4]. These waves were followed by a prolonged low-incidence phase dominated by the Delta (B.1.617.2) variant (Supplementary Figure S1) [5, 6]. The Alpha and Beta waves and the early phase of Delta incidence coincided with the rapid scale-up of coronavirus disease 2019 (COVID-19) vaccination using the BNT162b2 (Pfizer-BioNTech) [7] and mRNA-1273 (Moderna) [8] vaccines [9].

These dynamics provide an opportunity to investigate the interplay of the effects of previous infection and vaccination against symptomatic Alpha, Beta, and Delta infections. We therefore estimated protection of previous infection, of mRNA vaccination after the second dose and after the third/booster dose, and of hybrid immunity of previous infection and vaccination against infection with these variants as well as against any severe (acute-care hospitalization) [10], critical (intensive-care-unit hospitalization) [10], or fatal [11] COVID-19.

## METHODS

### Study population and data sources

This study was conducted on the population of Qatar between January 18, 2021, onset of the Alpha wave [3], through December 18, 2021, right before onset of the Omicron wave on December 19, 2021 [1]. The study analyzed the national, federated databases for COVID-19 laboratory testing, vaccination, hospitalization, and death, retrieved from the integrated, nationwide, digital-health information platform (Supplementary Text S1). Databases include all SARS-CoV-2-related data with no missing information since onset of the pandemic, including all polymerase chain reaction (PCR) tests regardless of location or facility (Supplementary Text S2). Every PCR test conducted in Qatar is classified on the basis of symptoms and the reason for testing. SARS-CoV-2 testing during the study was widely available and performed extensively, mostly for non-clinical reasons [5, 12]. Most infections were diagnosed not because of symptoms, but because of routine testing [5, 12]. Demographic information, such as sex and age, were extracted as registered in the national health registry. Further descriptions of Qatar’s population and of the national databases have been reported previously [5, 12–14].

### Study design

This study estimated effectiveness of previous pre-Omicron infection, vaccination with BNT162b2 or mRNA-1273, and hybrid immunity against symptomatic infection with Alpha, Beta, or Delta using a test-negative, case-control study design [1, 15–17]. The study design followed that developed earlier to investigate effects of previous infection, vaccination, and hybrid immunity against symptomatic Omicron infections [12]. This design estimates effectiveness by comparing odds of previous infection and/or vaccination among PCR-positive tests (cases) versus PCR-negative tests (controls) [1, 15–17].

In estimating effectiveness against symptomatic infection, we exactly matched cases and controls one-to-two by sex, 10-year age group, nationality, number of coexisting conditions (0, 1-2, or ≥3), and calendar week of PCR test. Matching was performed to balance observed confounders between exposure groups that are related to risk of infection in Qatar [18–22]. Choice of matching factors was informed by results of prior studies on Qatar’s population [5, 9, 23–25]. For estimating effectiveness against any severe [10], critical [10], or fatal [11] COVID-19, a one-to-five matching ratio was used to enhance statistical precision.

Only the first PCR-positive test occurring during a variant-dominated period was included for cases, while all PCR-negative tests were included for controls. Controls consisted of PCR-negative tests for individuals with no record of a PCR-positive test during that period. Only PCR tests conducted because of clinical suspicion due to presence of symptoms compatible with a respiratory tract infection were analyzed.

SARS-CoV-2 reinfection is conventionally defined as a documented infection ≥90 days after a previous infection, to avoid misclassification of prolonged PCR positivity as reinfection, if a shorter time interval is used [1, 26]. Previous infection was thus defined as a PCR-positive test ≥90 days before this study’s PCR test. Cases or controls with PCR-positive tests <90 days before the study’s PCR test were excluded.

Tests on individuals who received vaccines other than BNT162b2 or mRNA-1273, or who received mixed vaccines, were excluded. SARS-CoV-2 tests conducted for travel-related requirements were excluded [5, 25]. Tests occurring within 14 days of a second vaccine dose or 7 days of a third (booster) dose were excluded. These inclusion and exclusion criteria were implemented to allow for immunity build-up after vaccination [13, 27], and to minimize different types of potential bias, as informed by earlier analyses on the same population [5, 17, 25]. Every control that met the inclusion criteria and that could be matched to a case was included.

The study compared five exposure groups who had one or more immunological events of infection and/or vaccination to those with no previous infection and no vaccination. These groups included individuals with only previous infection, only two-dose (primary-series) vaccination, only three-dose (primary-series plus booster) vaccination, previous infection and two-dose vaccination, and previous infection and three-dose vaccination. These groups were defined based on status of prior immunological events at the time of the PCR test.

Classification of COVID-19 case severity [10], criticality [10], and fatality [11] followed World Health Organization guidelines, based on a national protocol applied to hospitalized COVID-19 patients (Supplementary Text S3).

### Variant ascertainment

The variant status of each infection was determined by the variant that dominated incidence at time of infection diagnosis. Duration of dominance of each variant was per Qatar’s variant genomic surveillance [28–30]. This surveillance consists of viral genome sequencing [28] and multiplex real-time reverse-transcription PCR (RT-qPCR) variant screening [29] of weekly collected random positive clinical samples, complemented by deep sequencing of wastewater samples [30] (Supplementary Text S2). Accordingly, an Alpha, Beta, or Delta infection was proxied as an infection diagnosed during January 18-March 7, 2021, March 8-May 31, 2021, or May 31-December 18, 2021, respectively.

### Statistical analysis

While all records of PCR testing were examined for selection of cases and controls, only matched samples were analyzed. Cases and controls were described using frequency distributions and measures of central tendency and compared using standardized mean differences (SMDs). An SMD of ≤0.1 indicated adequate matching [31].

Odds ratios, comparing odds of previous infection and/or vaccination among cases versus controls, and associated 95% confidence intervals (CIs) were derived using conditional logistic regression. This analytical approach, that also factors matching by calendar week of test, minimizes potential bias due to variation in epidemic phase and roll-out of vaccination during the study [15, 32]. CIs were not adjusted for multiplicity and interactions were not investigated. The reference group for all estimates comprised individuals with no previous infection and no vaccination. Effectiveness measures and associated 95% CIs were calculated as 1-odds ratio of previous infection and/or vaccination among cases versus controls (Supplementary Text S1).

Estimated effectiveness of hybrid immunity of previous infection and vaccination was compared to predicted effectiveness of hybrid immunity of previous infection and vaccination as calculated assuming that the effects of each of previous infection and vaccination act independently. If these two immunity effects are independent, effectiveness of hybrid immunity (*z*) is given by *z* = 1− (1− *x*)×(1− *y*), where *x* is effectivensss of previous infection alone and *y* is effecteivness of vaccination alone. This independence-model effectiveness was calculated against Alpha, Beta, and Delta as well as against any Omicron subvariant and against BA.1 and BA.2 Omicron subvariants. The Omicron estimates for previous infection and vaccination used as input in the independence-model equation were published earlier by Altarawneh et al. [12]. Statistical analyses were conducted in STATA/SE version 17.0 (Stata Corporation, College Station, TX, USA).

### Ethical approval

The institutional review boards at Hamad Medical Corporation and Weill Cornell Medicine– Qatar approved this retrospective study with a waiver of informed consent. The study was reported according to the Strengthening the Reporting of Observational Studies in Epidemiology (STROBE) guidelines (Supplementary Table S1).

## RESULTS

### Study population

Between December 23, 2020, date of first vaccination in Qatar [27], and December 18, 2021, end of study, 1,286,955 individuals received at least two BNT162b2 doses, of whom 152,316 received a third (booster) dose. The median date was May 2, 2021 for the first dose, May 23, 2021 for the second dose, and November 25, 2021 for the third dose. The median duration between the first and second doses was 21 days (interquartile range (IQR), 21-22 days), and the median duration between the second and third doses was 247 days (IQR, 238-258 days).

Meanwhile, 887,773 individuals received at least two mRNA-1273 doses, of whom 26,598 received a third dose. The median date was May 27, 2021 for the first dose, June 27, 2021 for the second dose, and December 6, 2021 for the third dose. The median duration between the first and second doses was 28 days (IQR, 28-30), and between the second and third doses was 216 days (IQR, 207-225).

Supplementary Figures S2 and S3 show the selection process of study populations for each of the BNT162b2 and mRNA-1273 analyses, respectively. Table 1 and Supplementary Table S2 describe the characteristics of study populations for these analyses, respectively.

**Table 1.**
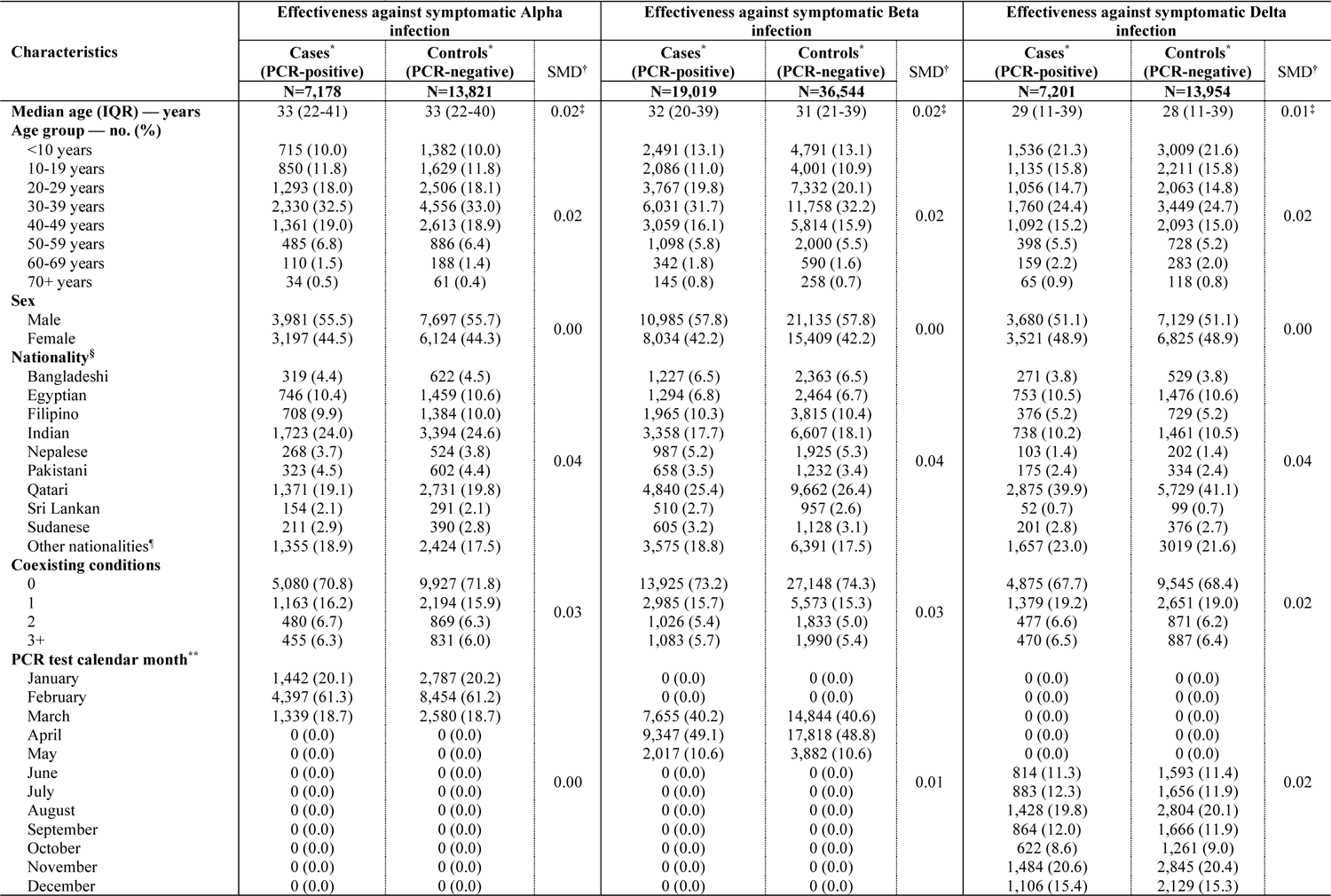
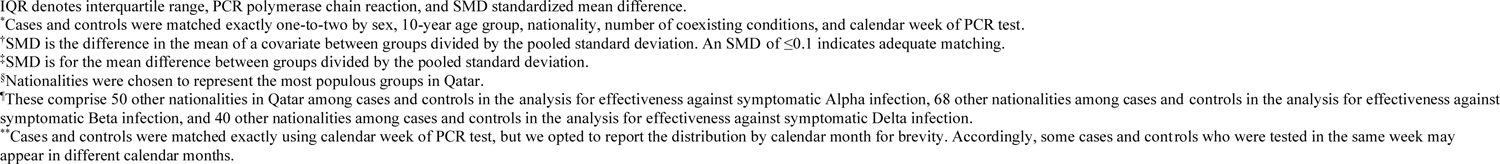
Characteristics of matched cases and controls in samples used to estimate effectiveness against symptomatic Alpha, Beta, or Delta infections in the BNT162b2 analysis.

### Effectiveness against symptomatic Alpha infection

#### BNT162b2 analysis

Effectiveness of only previous infection against symptomatic Alpha infection was 89.5% (95% CI: 85.5-92.3%) (Figure 1A and Table 2). The median duration between the previous infection and PCR test was 226.5 days (IQR, 169-257 days). Effectiveness of only two-dose BNT162b2 vaccination was 90.5% (95% CI, 83.9-94.4%). The median duration between the second dose and PCR test was 25 days (IQR, 19-32 days). Effectiveness of hybrid immunity of previous infection and two-dose BNT162b2 vaccination was 100.0% (95% CI: 84.1-100.0%).

**Figure 1.**
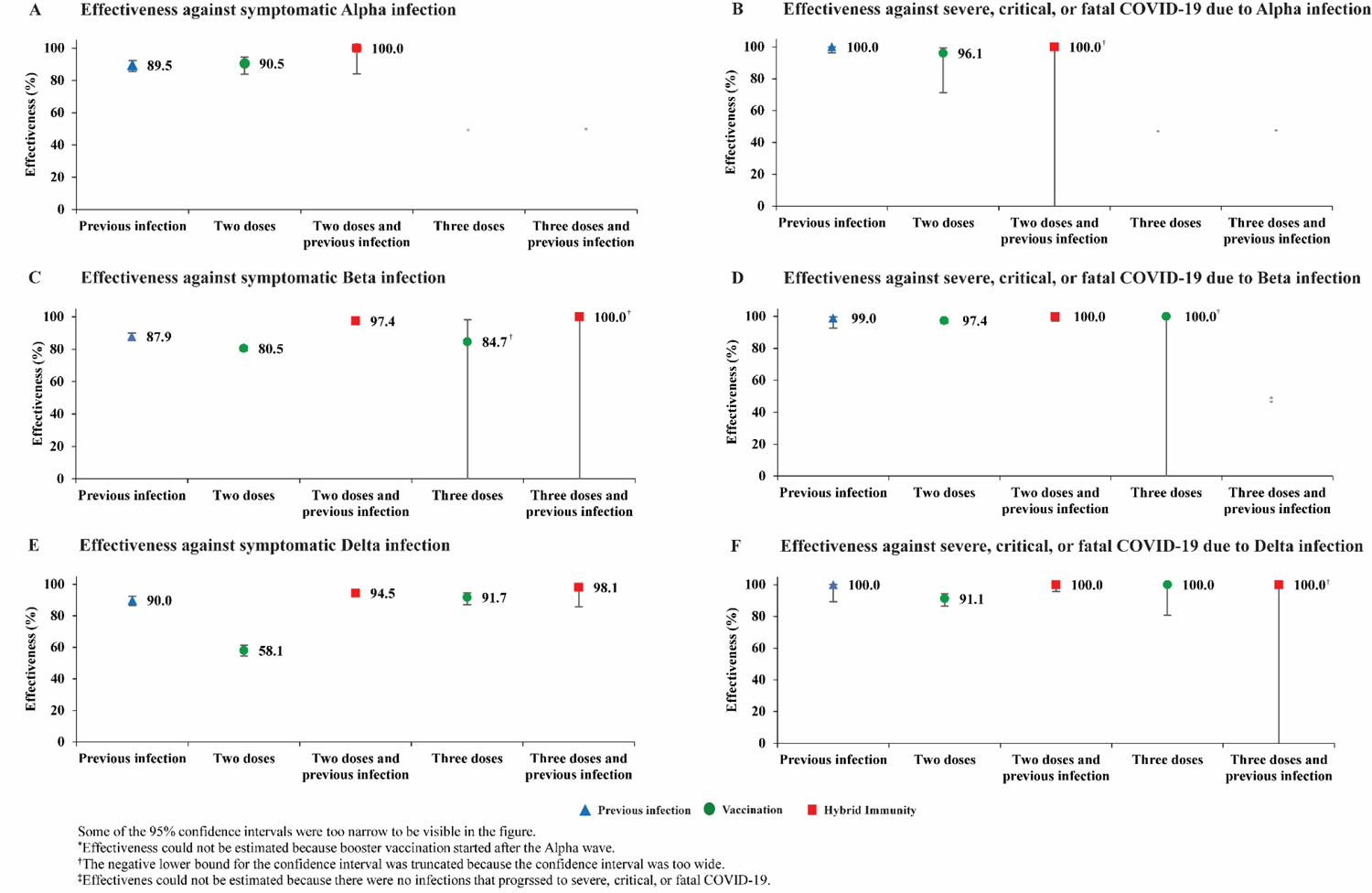
Effectiveness of previous pre-Omicron infection, vaccination with BNT162b2, and hybrid immunity of previous infection and vaccination against symptomatic Alpha, Beta, or Delta infections and against severe, critical, or fatal COVID-19 due to infection with these variants.

**Table 2.**
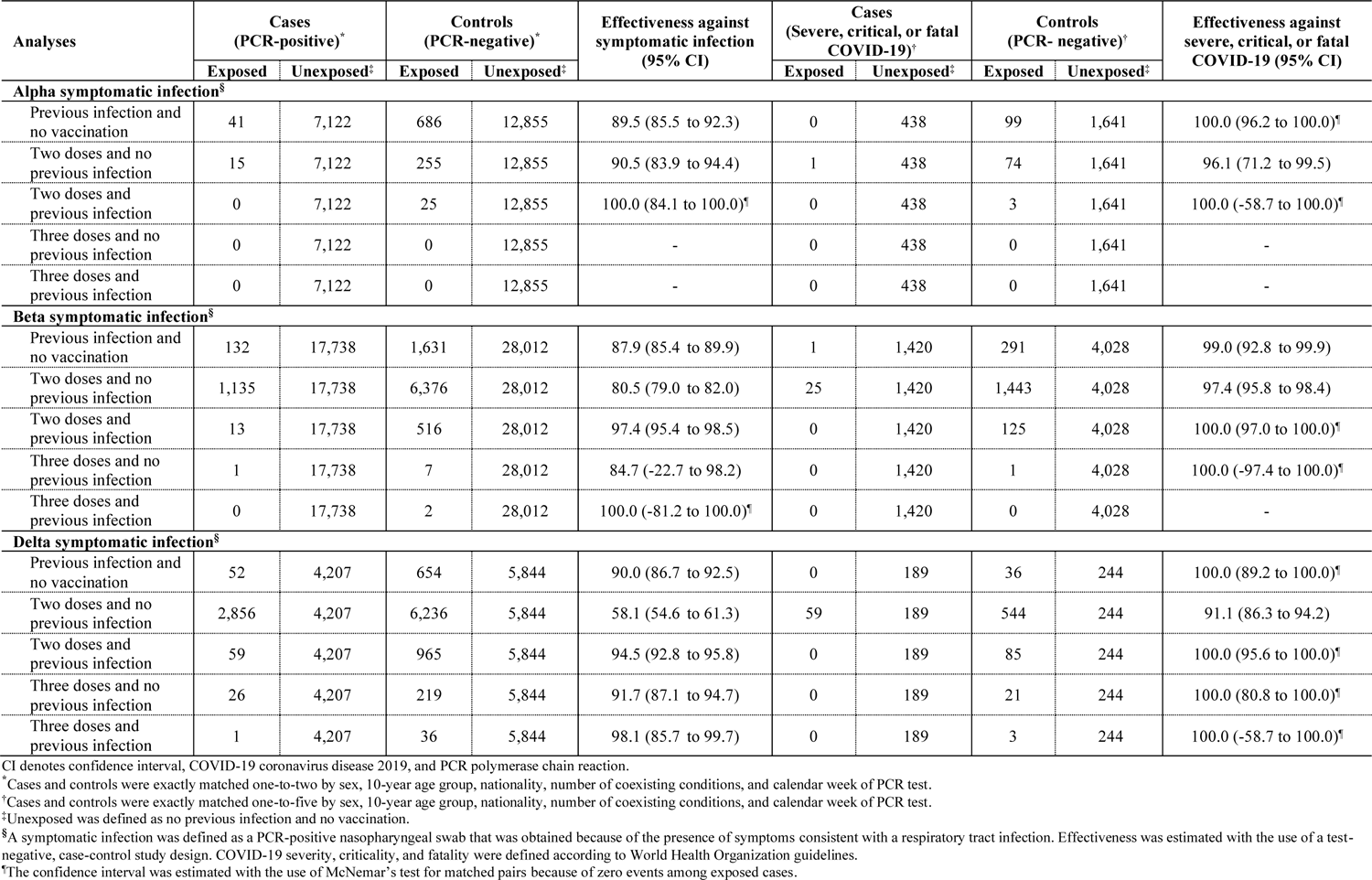
Effectiveness of previous pre-Omicron infection, vaccination with BNT162b2, and hybrid immunity of previous infection and vaccination against symptomatic Alpha, Beta, or Delta infections and against severe, critical, or fatal COVID-19 due to infection with these variants.

#### mRNA-1273 analysis

It was not possible to estimate mRNA-1273 effectiveness against Alpha, as number of mRNA-1273 vaccinations was limited. However, it was possible to provide another estimate for effectiveness of only previous infection against symptomatic Alpha infection at 88.3% (95% CI: 82.8-92.1%) (Figure 2A and Supplementary Table S3). The median duration between the previous infection and PCR test was 232 days (IQR, 184.5-261 days).

**Figure 2.**
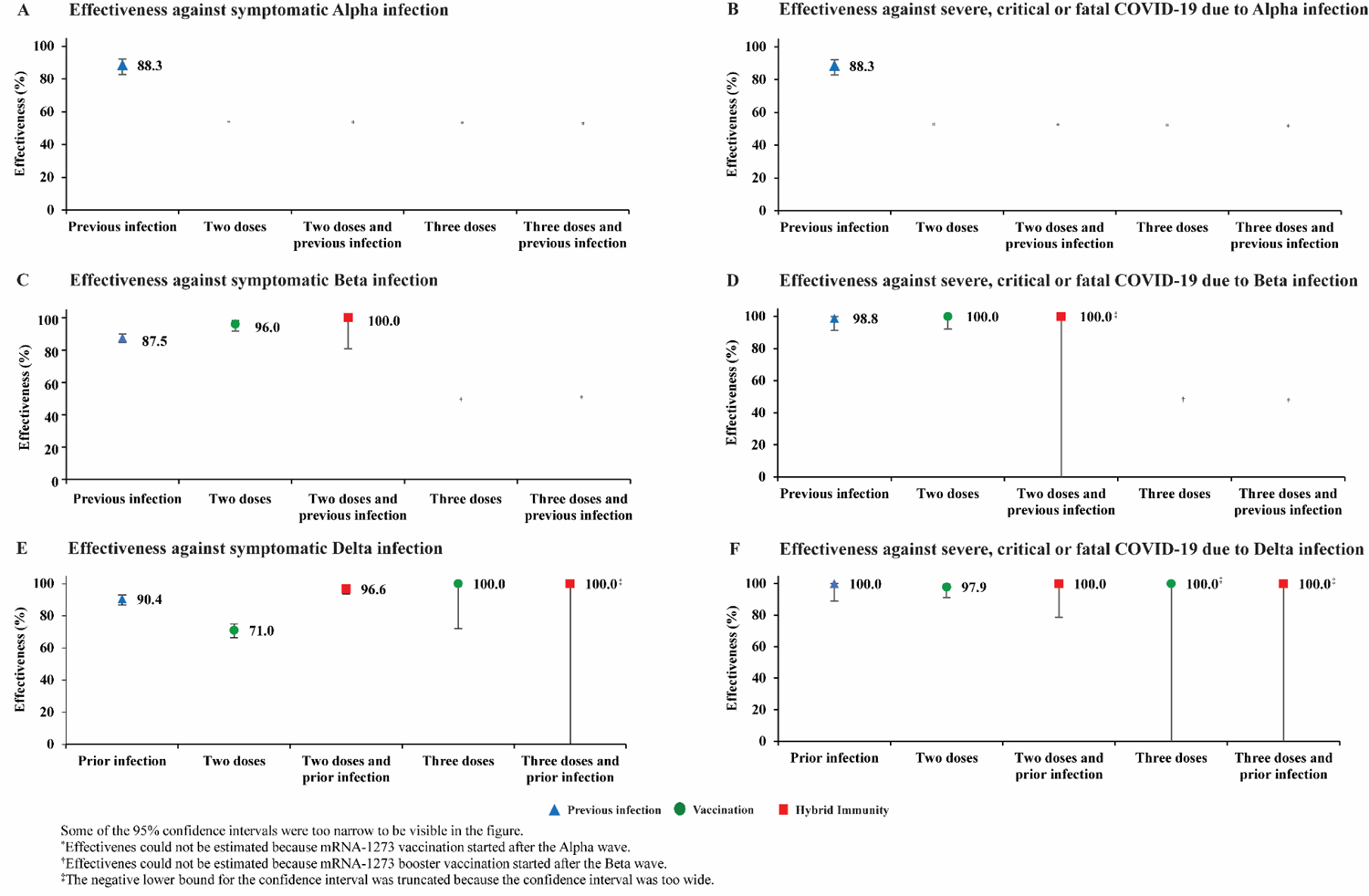
Effectiveness of previous pre-Omicron infection, vaccination with mRNA-1273, and hybrid immunity of previous infection and vaccination against symptomatic Alpha, Beta, or Delta infections and against severe, critical, or fatal COVID-19 due to infection with these variants.

### Effectiveness against symptomatic Beta infection

#### BNT162b2 analysis

Effectiveness of only previous infection against symptomatic Beta infection was 87.9% (95% CI: 85.4-89.9%) (Figure 1C and Table 2). The median duration between the previous infection and PCR test was 272 days (IQR, 213-307 days).

Effectiveness of only two-dose BNT162b2 vaccination was 80.5% (95% CI: 79.0-82.0%). The median duration between the second dose and PCR test was 35 days (IQR, 23-53 days).

Effectiveness of only three-dose BNT162b2 vaccination was 84.7% (95% CI: −22.7-98.2%). The median duration between the third dose and PCR test was 25 days (IQR, 21-34 days).

Effectiveness of hybrid immunity of previous infection and two-dose BNT162b2 vaccination was 97.4% (95% CI: 95.4-98.5%). Effectiveness of previous infection and three-dose BNT162b2 vaccination was 100.0% (95% CI: −81.2-100.0%).

#### mRNA-1273 analysis

Effectiveness of only previous infection against symptomatic Beta infection was 87.5% (95% CI: 84.6-89.9%) (Figure 2C and Supplementary Table S3). The median duration between the previous infection and PCR test was 277 days (IQR, 214-308 days). Effectiveness of only two-dose mRNA-1273 vaccination was 96.0% (95% CI, 91.6-98.1%). The median duration between the second dose and PCR test was 21 days (IQR, 17-30 days). Effectiveness of hybrid immunity of previous infection and two-dose mRNA-1273 vaccination was 100.0% (95% CI: 80.8-100.0%).

### Effectiveness against symptomatic Delta infection

#### BNT162b2 analysis

Effectiveness of only previous infection against symptomatic Delta infection was 90.0% (95% CI: 86.7-92.5%) (Figure 1E and Table 2). The median duration between the previous infection and PCR test was 280 days (IQR, 196-404 days).

Effectiveness of only two-dose BNT162b2 vaccination was 58.1% (95% CI: 54.6-61.3%). The median duration between the second dose and PCR test was 150 days (IQR, 98-204 days).

Effectiveness of only three-dose BNT162b2 vaccination was 91.7% (95% CI: 87.1-94.7%). The median duration between the third dose and PCR test was 23.5 days (IQR, 14-43 days).

Effectiveness of hybrid immunity of previous infection and two-dose BNT162b2 vaccination was 94.5% (95% CI: 92.8-95.8%). Effectiveness of previous infection and three-dose BNT162b2 vaccination was 98.1% (95% CI: 85.7-99.7%).

#### mRNA-1273 analysis

Effectiveness of only previous infection against symptomatic Delta infection was 90.4% (95% CI: 86.8-93.0%) (Figure 2E and Supplementary Table S3). The median duration between the previous infection and PCR test was 258.5 days (IQR, 186-389 days).

Effectiveness of only two-dose mRNA-1273 vaccination was 71.0% (95% CI: 66.4-74.9%). The median duration between the second dose and PCR test was 111 days (IQR, 59-168 days).

Effectiveness of only three-dose mRNA-1273 vaccination was 100.0% (95% CI: 72.1-100.0%). The median duration between the third dose and PCR test was 19 days (IQR, 12-30 days).

Effectiveness of hybrid immunity of previous infection and two-dose mRNA-1273 vaccination was 96.6% (95% CI: 93.5-98.2%). Effectiveness of previous infection and three-dose mRNA-1273 vaccination was 100.0% (95% CI: −8.4-100.0%).

### Effectiveness against severe, critical, or fatal COVID-19

Previous infection, vaccination, and hybrid immunity all showed robust effectiveness (>90%) against severe, critical, or fatal COVID-19 regardless of the underlying variant, but some of the 95% CIs were wide because of small case numbers (Figures 1 and 2, Table 2 and Supplementary Table S3).

### Hybrid immunity protection: Directly estimated versus independence-model prediction

Figure 3 shows the directly estimated effectiveness of hybrid immunity of previous infection and vaccination compared to that predicted by the independence model, that is assuming independence of the effects of previous-infection and vaccination. Both estimates were comparable (Pearson correlation: 0.99).

**Figure 3.**
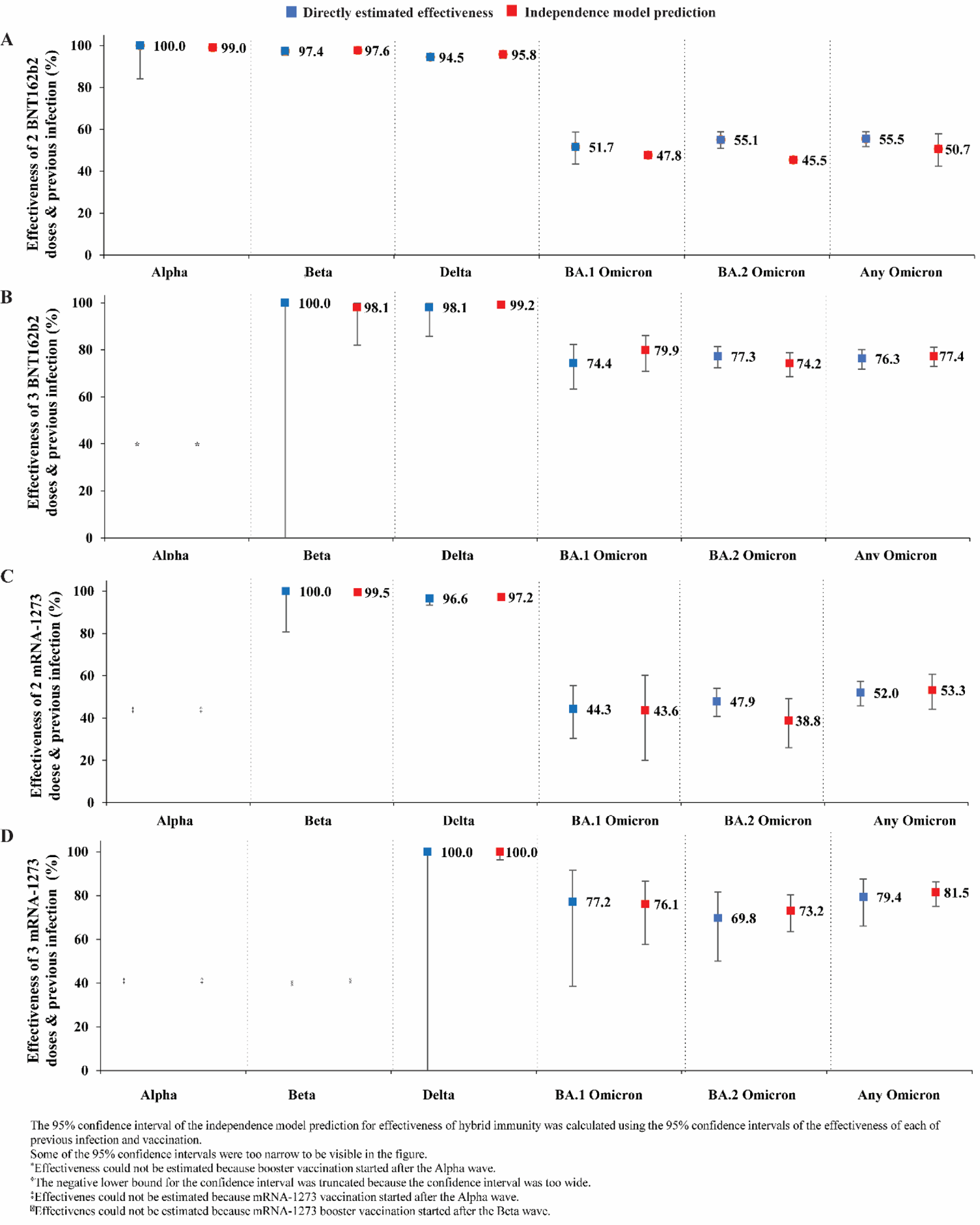
Estimated effectiveness of hybrid immunity against symptomatic Alpha, Beta, Delta, BA.1 Omicron, BA.2 Omicron, and any Omicron infection compared to predicted effectiveness against these infections assuming independence of the effects of previous infection immunity and vaccination immunity (independence model prediction).

## DISCUSSION

Hybrid immunity of natural infection and vaccination provided higher protection against infection than each of natural infection or vaccination alone, regardless of variant. This finding is consistent with epidemiological and laboratory studies indicating superior protection for hybrid immunity [12, 33, 34]. Strikingly, the level of protection of hybrid immunity was similar to that predicted from the individual protections of previous infection and vaccination by assuming that each acted independently of the other one. The combined effect of these two forms of protection reflected neither synergy nor redundancy of their individual biological effects. This was true regardless of variant, not only against Alpha, Beta, and Delta, but also against Omicron subvariants, as suggested earlier [12]. This finding, which may only apply in the first few months after infection or vaccination [14, 35], is perhaps a consequence of neutralizing antibodies being behind the protection against infection [36]. Each of infection and vaccination elicits independently production of antibody titers, and the combined effect of both titers results in a stronger protection for hybrid immunity against infection.

Protection of a pre-Omicron infection against reinfection with Alpha, Beta, or Delta was strong at ∼90%. This finding confirms the series of studies indicating strong protection for pre-Omicron infection against pre-Omicron reinfection, including on this same population [1-4, 17, 26, 37, 38]. Protection of primary-series vaccination against Alpha or Beta was also strong at >80% for both BNT162b2 and mRNA-1273, confirming earlier findings [24, 27]. Protection against Delta was lower, at ∼60% for BNT162b2 and at ∼70% for mRNA-1273, supporting earlier findings [6], but also reflecting the waning of primary-series vaccine protection by time of Delta dominance [5, 6, 25]. Protection of the booster dose against Delta was strong at >90% for both vaccines, a consequence of the booster dose being recent, also supporting earlier findings [13].

While there were differences in the protection of previous infection, vaccination, and hybrid immunity against infection, all of these forms of immunity had very strong protection against severe, critical, or fatal COVID-19, irrespective of variant, at >90%. This finding affirms the strong protection of any form of immunity against severe infection and that breakthrough infections, when they occur, are not likely to be severe. This finding is consistent with other findings indicating that reinfections are ∼90% less likely to be severe than primary infections [37, 39], and that vaccination induces strong protection against severe COVID-19 that lasts longer than the vaccine’s protection against infection [5, 6, 25].

This study has limitations. With the relatively young population of Qatar [18], our findings may not be generalizable to other countries where elderly citizens constitute a large proportion of the population. The study used an observational test-negative design, but bias can arise in such design in unexpected ways, or from unknown sources, such as subtle differences in test-seeking behavior or changes in the pattern of testing. Variant ascertainment was based on a time criteria of the variant that dominated incidence and not based on viral genome sequencing or genotyping of every infection.

While matching was done for several factors, it was not possible for other factors such as geography or occupation, as such data were unavailable. However, Qatar is essentially a city state and infection incidence was distributed across neighborhoods. Nationality, age, and sex provide a powerful proxy for socio-economic status in this country [18–22], and thus matching by these factors may have also (partially) controlled for other factors such as occupation. This matching prescription had already been investigated in previous studies of different epidemiologic designs, and using control groups to test for null effects [5, 9, 23–25]. These studies have supported that this prescription provides adequate control of the differences in infection exposure [5, 9, 23–25]. The study was implemented on Qatar’s total population at a time of mass-scale PCR testing [5], perhaps minimizing the likelihood of bias.

Notwithstanding these limitations, findings are consistent with those of earlier studies that used different epidemiologic study designs on the same population [1, 3–6, 13, 17, 24, 25, 27, 37, 39]. Extensive sensitivity and additional analyses were conducted to investigate effects of potential bias in our earlier studies that used a similar methodology [5, 17, 25]. These included different adjustments in the analysis and various study inclusion and exclusion criteria, to investigate whether effectiveness estimates could have been biased [5, 17, 25]. These analyses confirmed the study findings [1, 5, 17, 25, 40].

In conclusion, all forms of natural and vaccine immunity prior to Omicron introduction provided strong protection against Alpha, Beta, and Delta infections, and very strong protection against severe COVID-19. Hybrid immunity of natural infection and vaccination provided higher protection against infection than that of natural infection or vaccination alone, regardless of variant. Level of hybrid-immunity protection was consistent with the individual protections of previous infection and vaccination acting independently of each other. This finding, however, may only apply in the first few months after infection or vaccination, when protection against infection is mediated by neutralizing antibodies.

## Data Availability

The dataset of this study is a property of the Qatar Ministry of Public Health that was provided to the researchers through a restricted-access agreement that prevents sharing the dataset with a third party or publicly. The data are available under restricted access for preservation of confidentiality of patient data. Access can be obtained through a direct application for data access to Her Excellency the Minister of Public Health (https://www.moph.gov.qa/english/OurServices/eservices/Pages/Governmental-HealthCommunication-Center.aspx). The raw data are protected and are not available due to data privacy laws. Aggregate data are available within the paper and its supplementary information.

## Acknowledgments

We acknowledge the many dedicated individuals at Hamad Medical Corporation, the Ministry of Public Health, the Primary Health Care Corporation, Qatar Biobank, Sidra Medicine, and Weill Cornell Medicine-Qatar for their diligent efforts and contributions to make this study possible.

## Author contributions

HNA co-designed the study, performed the statistical analyses, and co-wrote the first draft of the article. HC co-designed the study and co-led the statistical analyses. LJA conceived and co-designed the study, co-led the statistical analyses, and co-wrote the first draft of the article. HNA, HC, and LJA accessed and verified all the data. PT and MRH designed and conducted multiplex, RT-qPCR variant screening and viral genome sequencing. PVC designed mass PCR testing to allow routine capture of SGTF variants and conducted viral genome sequencing. HY, HAK, and MS conducted viral genome sequencing. All authors contributed to data collection and acquisition, database development, discussion and interpretation of the results, and to the writing of the article. All authors have read and approved the final manuscript.

## Supplementary Material

### Supplementary Text S1. Further details on methods

#### Data sources and testing

Qatar’s national and universal public healthcare system uses the Cerner-system advanced digital health platform to track all electronic health record encounters of each individual in the country, including all citizens and residents registered in the national and universal public healthcare system. Registration in the public healthcare system is mandatory for citizens and residents.

The databases analyzed in this study are data-extract downloads from the Cerner-system that have been implemented on a regular (twice weekly) schedule since the onset of pandemic by the Business Intelligence Unit at Hamad Medical Corporation. Hamad Medical Corporation is the national public healthcare provider in Qatar. At every download all tests, coronavirus disease 2019 (COVID-19) vaccinations, hospitalizations related to COVID-19, and all death records regardless of cause are provided to the authors through .csv files. These databases have been analyzed throughout the pandemic not only for study-related purposes, but also to provide policymakers with summary data and analytics to inform the national response.

Every health encounter in the Cerner-system is linked to a unique individual through the HMC Number that links all records for this individual at the national level. Databases were merged and analyzed using the HMC Number to link all records whether for testing, vaccinations, hospitalizations, and deaths. All deaths in Qatar are tracked by the public healthcare system. All COVID-19-related healthcare was provided only in the public healthcare system. No private entity was permitted to provide COVID-19-related healthcare. COVID-19 vaccination was also provided only through the public healthcare system. These health records were tracked throughout the COVID-19 pandemic using the Cerner system. This system has been implemented in 2013, before the onset of the pandemic. Therefore, we had the health records related to this study for the full national cohort of citizens and residents throughout the pandemic.

Demographic details for every HMC Number (individual) such as sex, age, and nationality are collected upon issuing of the universal health card, based on the Qatar Identity Card, which is a mandatory requirement by the Ministry of Interior to every citizen and resident in the country.

Severe acute respiratory syndrome coronavirus 2 (SARS-CoV-2) testing in Qatar is done at a mass scale where close to 5% of the population are tested every week [1, 2]. All SARS-CoV-2 testing in any facility in this country is tracked nationally in one database, the national testing database. This database covers all testing in all locations and facilities throughout the country, whether public or private. Every polymerase chain reaction (PCR) test, regardless of location or setting, is classified on the basis of symptoms and the reason for testing (clinical symptoms, contact tracing, surveys or random testing campaigns, individual requests, routine healthcare testing, pre-travel, at port of entry, or other). Based on the distribution of the reason for testing, most of the tests that have been conducted in Qatar were conducted for routine reasons, such as being travel-related. About 75% of those diagnosed are also diagnosed not because of appearance of symptoms, but because of routine testing [1, 2].

Qatar has unusually young, diverse demographics, in that only 9% of its residents are ≥50 years of age, and 89% are expatriates from over 150 countries [3, 4]. Further descriptions of the study population and these national databases were reported previously [1, 2, 4–7].

#### Comorbidity classification

Comorbidities were ascertained and classified based on the ICD-10 codes for chronic conditions as recorded in the electronic health record encounters of each individual in the Cerner-system national database that includes all citizens and residents registered in the national and universal public healthcare system. The public healthcare system provides healthcare to the entire resident population of Qatar free of charge or at heavily subsidized costs, including prescription drugs. With the mass expansion of this sector in recent years, facilities have been built to cater to specific needs of subpopulations. For example, tens of facilities have been built, including clinics and hospitals, in localities with high density of craft and manual workers [8].

All encounters for each individual were analyzed to determine the comorbidity classification for that individual, as part of a recent national analysis to assess healthcare needs and resource allocation. The Cerner-system national database includes encounters starting from 2013, after this system was launched in Qatar. As long as each individual had at least one encounter with a specific comorbidity diagnosis since 2013, this person was classified with this comorbidity. Individuals who have comorbidities but never sought care in the public healthcare system, or seek care exclusively in private healthcare facilities, were classified as individuals with no comorbidity due to absence of recorded encounters for them.

#### Calculation of previous infection and vaccination effectiveness

Effectiveness measures and associated 95% CIs were calculated as 1-odds ratio (OR) of previous infection and/or vaccination among cases versus controls if the OR<1, and as 1/OR-1 if the OR was ≥1 [9–11]. The latter was done to ensure symmetric scale for both negative and positive effectiveness, ranging from −100%-100%, leading to easier and meaningful interpretation of effectiveness, regardless of being positive or negative.

### Supplementary Text S2. Laboratory methods and variant ascertainment

#### Real-time reverse-transcription polymerase chain reaction testing

Nasopharyngeal and/or oropharyngeal swabs were collected for polymerase chain reaction (PCR) testing and placed in Universal Transport Medium (UTM). Aliquots of UTM were: 1) extracted on KingFisher Flex (Thermo Fisher Scientific, USA), MGISP-960 (MGI, China), or ExiPrep 96 Lite (Bioneer, South Korea) followed by testing with real-time reverse-transcription PCR (RT-qPCR) using TaqPath COVID-19 Combo Kits (Thermo Fisher Scientific, USA) on an ABI 7500 FAST (Thermo Fisher Scientific, USA); 2) tested directly on the Cepheid GeneXpert system using the Xpert Xpress SARS-CoV-2 (Cepheid, USA); or 3) loaded directly into a Roche cobas 6800 system and assayed with the cobas SARS-CoV-2 Test (Roche, Switzerland). The first assay targets the viral S, N, and ORF1ab gene regions. The second targets the viral N and E-gene regions, and the third targets the ORF1ab and E-gene regions. All PCR testing was conducted at the Hamad Medical Corporation Central Laboratory or Sidra Medicine Laboratory, following standardized protocols.

#### Classification of infections by variant type

Surveillance for the severe acute respiratory syndrome coronavirus 2 (SARS-CoV-2) variants in Qatar is based on viral genome sequencing and multiplex real-time reverse-transcription polymerase chain reaction (RT-qPCR) variant screening [12] of weekly collected random positive clinical samples [2, 13–17], complemented by deep sequencing of wastewater samples [15, 18, 19]. Further details on the viral genome sequencing and multiplex RT-qPCR variant screening throughout the SARS-CoV-2 waves in Qatar can be found in previous publications [1, 2, 6, 13–17, 20–25].

### Supplementary Text S3. COVID-19 severity, criticality, and fatality classification

Classification of Coronavirus Disease 2019 (COVID-19) case severity (acute-care hospitalizations) [26], criticality (intensive-care-unit hospitalizations) [26], and fatality [27] followed World Health Organization (WHO) guidelines. Assessments were made by trained medical personnel independent of study investigators and using individual chart reviews, as part of a national protocol applied to every hospitalized COVID-19 patient. Each hospitalized COVID-19 patient underwent an infection severity assessment every three days until discharge or death. We classified individuals who progressed to severe, critical, or fatal COVID-19 between the time of the documented infection and the end of the study based on their worst outcome, starting with death [27], followed by critical disease [26], and then severe disease [26].

Severe COVID-19 disease was defined per WHO classification as a SARS-CoV-2 infected person with “oxygen saturation of <90% on room air, and/or respiratory rate of >30 breaths/minute in adults and children >5 years old (or ≥60 breaths/minute in children <2 months old or ≥50 breaths/minute in children 2-11 months old or ≥40 breaths/minute in children 1–5 years old), and/or signs of severe respiratory distress (accessory muscle use and inability to complete full sentences, and, in children, very severe chest wall indrawing, grunting, central cyanosis, or presence of any other general danger signs)” [26]. Detailed WHO criteria for classifying Severe acute respiratory syndrome coronavirus 2 (SARS-CoV-2) infection severity can be found in the WHO technical report [26].

Critical COVID-19 disease was defined per WHO classification as a SARS-CoV-2 infected person with “acute respiratory distress syndrome, sepsis, septic shock, or other conditions that would normally require the provision of life sustaining therapies such as mechanical ventilation (invasive or non-invasive) or vasopressor therapy” [26]. Detailed WHO criteria for classifying SARS-CoV-2 infection criticality can be found in the WHO technical report [26].

COVID-19 death was defined per WHO classification as “a death resulting from a clinically compatible illness, in a probable or confirmed COVID-19 case, unless there is a clear alternative cause of death that cannot be related to COVID-19 disease (e.g. trauma). There should be no period of complete recovery from COVID-19 between illness and death. A death due to COVID-19 may not be attributed to another disease (e.g. cancer) and should be counted independently of preexisting conditions that are suspected of triggering a severe course of COVID-19”. Detailed WHO criteria for classifying COVID-19 death can be found in the WHO technical report [27].

---

**Supplementary Table S1.** Strengthening the Reporting of Observational Studies in Epidemiology (STROBE) checklist for case-control studies.

|  | Item No | Recommendation | Main text page |
| --- | --- | --- | --- |
| Title and abstract | 1 | (a) Indicate the study’s design with a commonly used term in the title or the abstract | Abstract |
|  |  | (b) Provide in the abstract an informative and balanced summary of what was done and what was found | Abstract |
| Introduction |  |  |  |
| Background/rationale | 2 | Explain the scientific background and rationale for the investigation being reported | Introduction |
| Objectives | 3 | State specific objectives, including any prespecified hypotheses | Introduction |
| Methods |  |  |  |
| Study design | 4 | Present key elements of study design | Methods (‘Study design’) |
| Setting | 5 | Describe the setting, locations, and relevant dates, including periods of recruitment, exposure, follow-up, and data collection | Methods (‘Study population and data sources’, ‘Study design’, & ‘Variant ascertainment’) & Supplementary Texts S1, S2, & S3 |
| Participants | 6 | (a) Give the eligibility criteria, and the sources and methods of case ascertainment and control selection. Give the rationale for the choice of cases and controls<br>(b) For matched studies, give matching criteria and the number of controls per case | Methods (‘Study population and data sources’ & ‘Study design’), Supplementary Figures S2 & S3 |
| Variables | 7 | Clearly define all outcomes, exposures, predictors, potential confounders, and effect modifiers. Give diagnostic criteria, if applicable | Methods (‘Study design’, ‘Variant ascertainment’, & ‘Statistical Analysis’) & Supplementary Texts S1, S2, & S3 |
| Data sources/measurement | 8 | For each variable of interest, give sources of data and details of methods of assessment (measurement). Describe comparability of assessment methods if there is more than one group | Methods (‘Study population and data sources’, ‘Study design’, ‘Variant ascertainment’, & ‘Statistical analysis’) & Supplementary Texts S1, S2, & S3 |
| Bias | 9 | Describe any efforts to address potential sources of bias | Methods (‘Study design’ & ‘Statistical analysis’) |
| Study size | 10 | Explain how the study size was arrived at | Supplementary Figures S2 & S3 |
| Quantitative variables | 11 | Explain how quantitative variables were handled in the analyses. If applicable, describe which groupings were chosen and why | Methods (‘Study design’ & ‘Statistical analysis’), Table 1, & Supplementary Table S2 |
| Statistical methods | 12 | (a) Describe all statistical methods, including those used to control for confounding | Methods (‘Statistical analysis’) |
|  |  | (b) Describe any methods used to examine subgroups and interactions | Methods (‘Statistical analysis’, paragraph 2) |
|  |  | (c) Explain how missing data were addressed | Not applicable, see Methods (‘Study population and data sources’) |
|  |  | (d) If applicable, explain how matching of cases and controls was addressed | Methods (‘Study design’, paragraph 2 & ‘Statistical analysis’, paragraph 2) |
|  |  | (e) Describe any sensitivity analyses | Not applicable |
| Results |  |  |  |
| Participants | 13 | (a) Report numbers of individuals at each stage of study—eg numbers potentially eligible, examined for eligibility, confirmed eligible, included in the study, completing follow-up, and analysed<br>(b) Give reasons for non-participation at each stage<br>(c) Consider use of a flow diagram | Supplementary Figures S2 & S3 |
| Descriptive data | 14 | (a) Give characteristics of study participants (eg demographic, clinical, social) and information on exposures and potential confounders | Results (‘Study population’), Table 1 & Supplementary Table S2 |
|  |  | (b) Indicate number of participants with missing data for each variable of interest | Not applicable, see Methods (‘Study population and data sources’) |
| Outcome data | 15 | Report numbers in each exposure category, or summary measures of exposure | Results (‘Effectiveness against symptomatic Alpha infection’, ‘Effectiveness against symptomatic Beta infection’, ‘Effectiveness against symptomatic Delta infection’, ‘Effectiveness against severe, critical, or fatal COVID-19’, & ‘Hybrid immunity protection: Directly estimated versus independence-model prediction’), Figures 1, 2, 3, Table 2, & Supplementary Table S3 |
| Main results | 16 | (a) Give unadjusted estimates and, if applicable, confounder-adjusted estimates and their precision (eg, 95% confidence interval). Make clear which confounders were adjusted for and why they were included | Results ‘(‘Effectiveness against symptomatic Alpha infection’, ‘Effectiveness against symptomatic Beta infection’, ‘Effectiveness against symptomatic Delta infection’, ‘Effectiveness against severe, critical, or fatal COVID-19’, & ‘Hybrid immunity protection: Directly estimated versus independence-model prediction’), Figures 1, 2, 3, Table 2, & Supplementary Table S3 |
|  |  | (b) Report category boundaries when continuous variables were categorized | Table 1 & Supplementary Table S2 |
|  |  | (c) If relevant, consider translating estimates of relative risk into absolute risk for a meaningful time period | Not applicable |
| Other analyses | 17 | Report other analyses done—eg analyses of subgroups and interactions, and sensitivity analyses | Not applicable |
| **Discussion** |  |  |  |
| Key results | 18 | Summarise key results with reference to study objectives | Discussion, paragraphs 1-3 |
| Limitations | 19 | Discuss limitations of the study, taking into account sources of potential bias or imprecision. Discuss both direction and magnitude of any potential bias | Discussion, paragraphs 4-6 |
| Interpretation | 20 | Give a cautious overall interpretation of results considering objectives, limitations, multiplicity of analyses, results from similar studies, and other relevant evidence | Discussion, paragraph 7 |
| Generalisability | 21 | Discuss the generalisability (external validity) of the study results | Discussion, paragraphs 4-5 |
| **Other information** |  |  |  |
| Funding | 22 | Give the source of funding and the role of the funders for the present study and, if applicable, for the original study on which the present article is based | Funding |

**Supplementary Figure S1.**
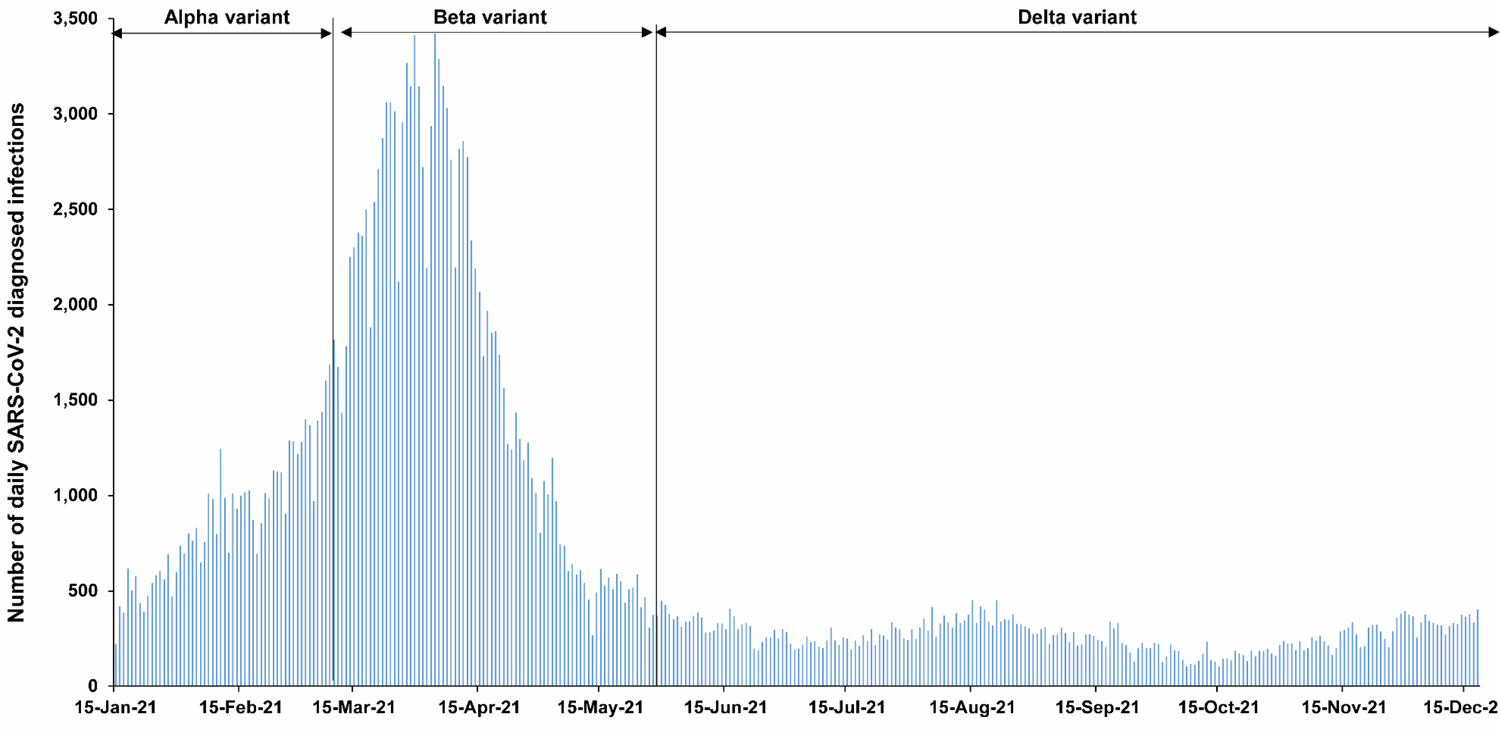
Daily number of newly diagnosed SARS-CoV-2 infections between January 15, 2021 and December 18, 2021.

**Supplementary Figure S2.**
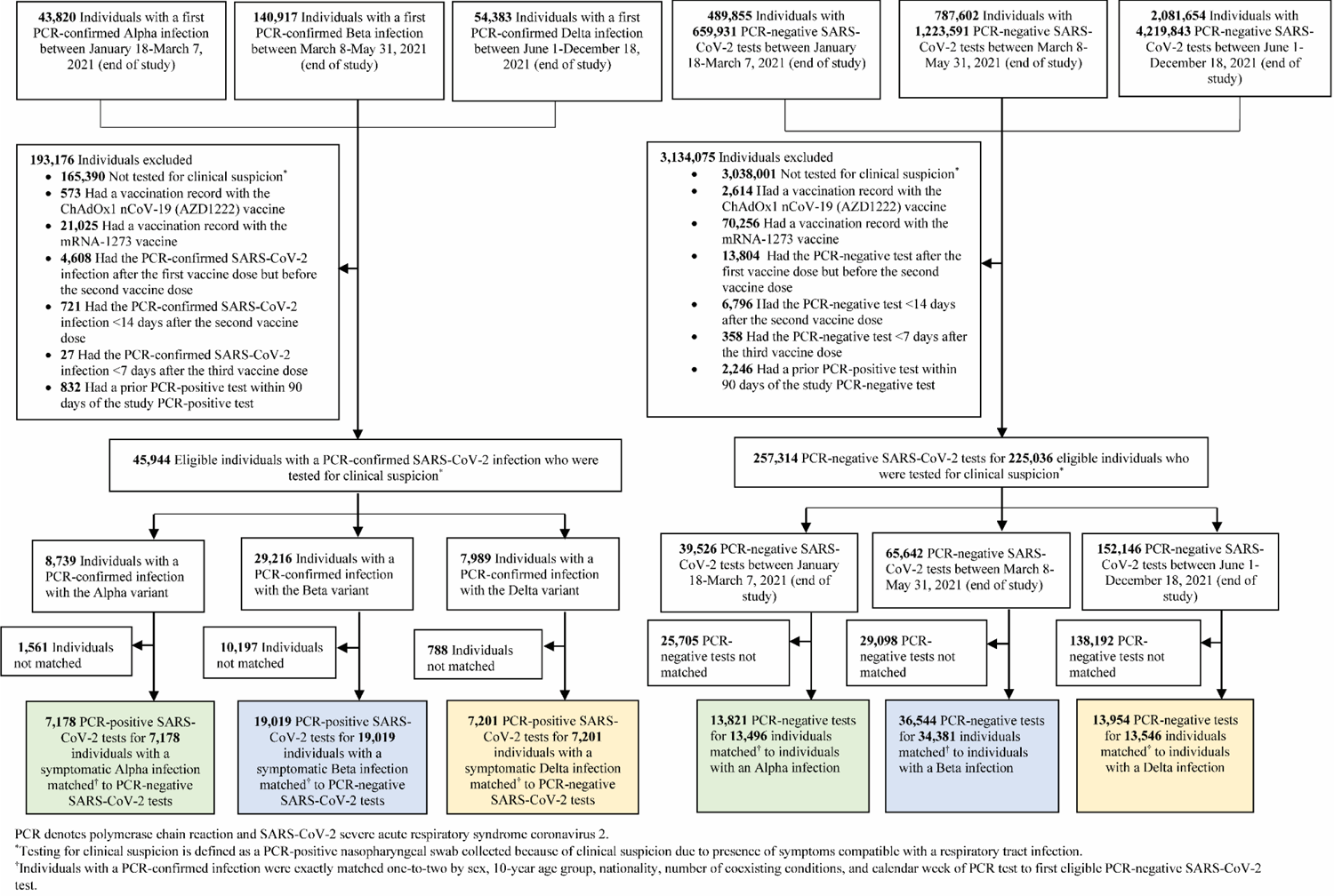
Flowchart describing the population selection process for investigating effectiveness of previous pre-Omicron infection, vaccination with BNT162b2, and hybrid immunity against symptomatic Alpha, Beta, or Delta infections.

**Supplementary Figure S3.**
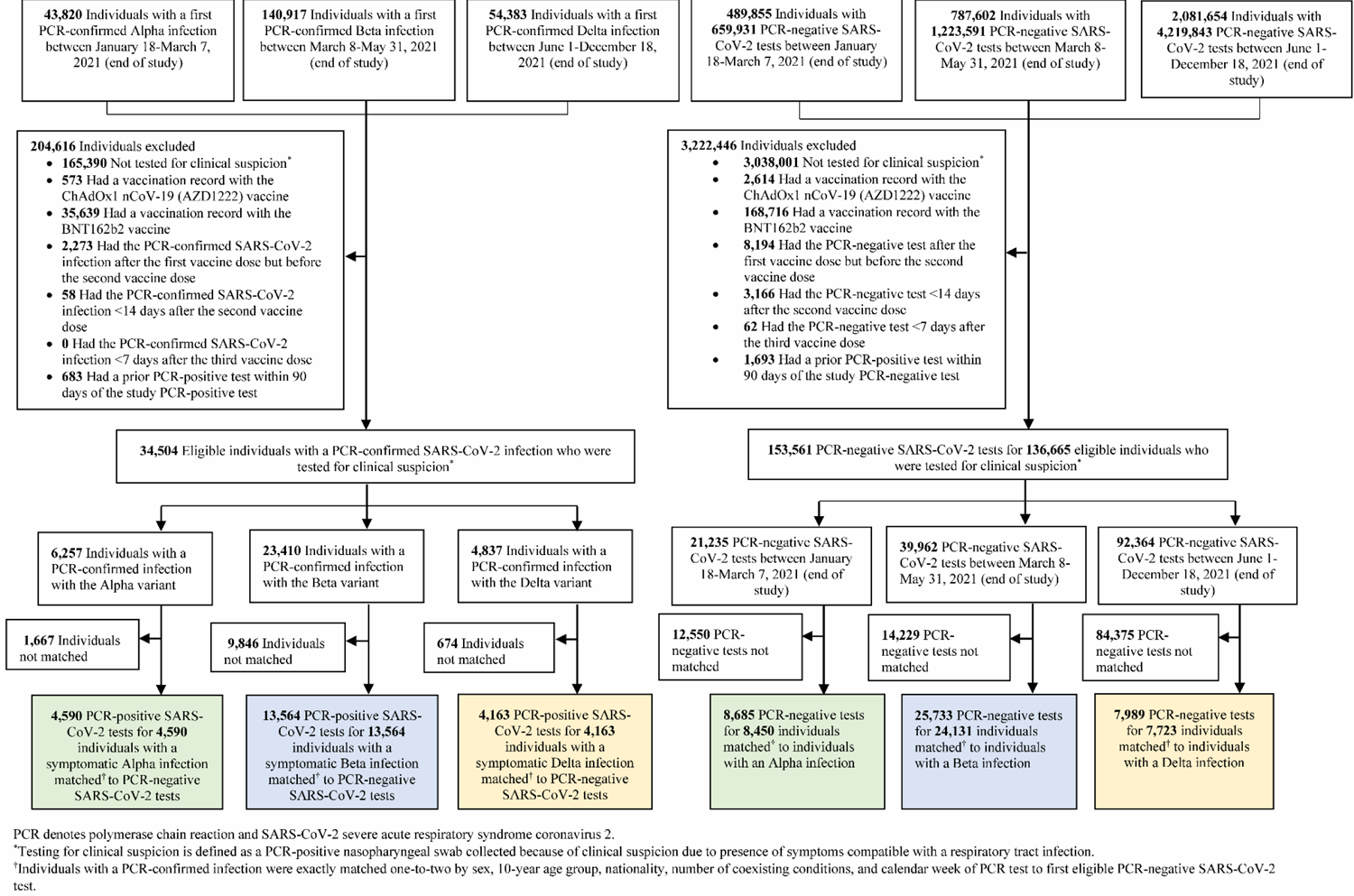
Flowchart describing the population selection process for investigating effectiveness of previous pre-Omicron infection, vaccination with mRNA-1273, and hybrid immunity against symptomatic Alpha, Beta, or Delta infections.

**Supplementary Table S2.**
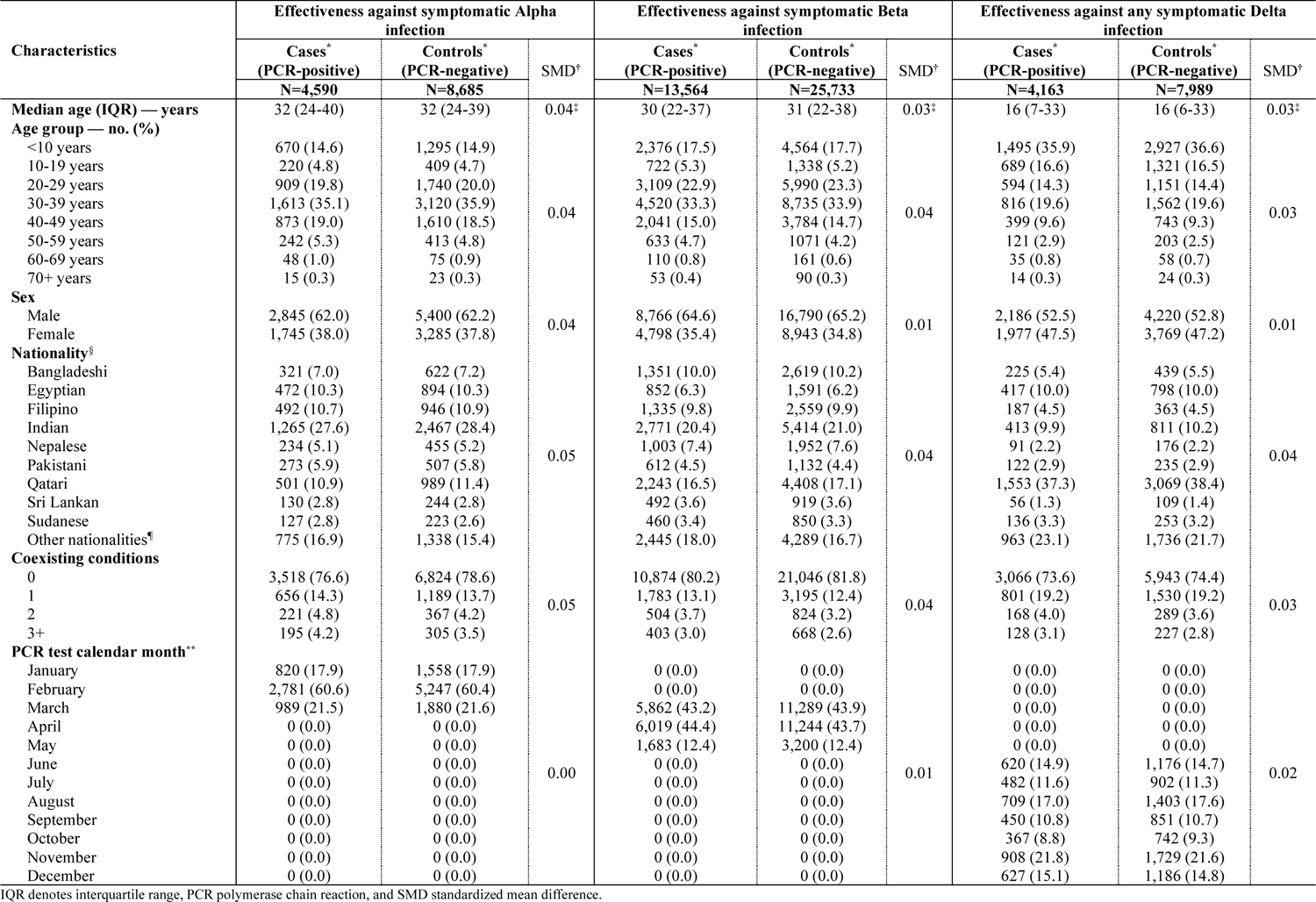
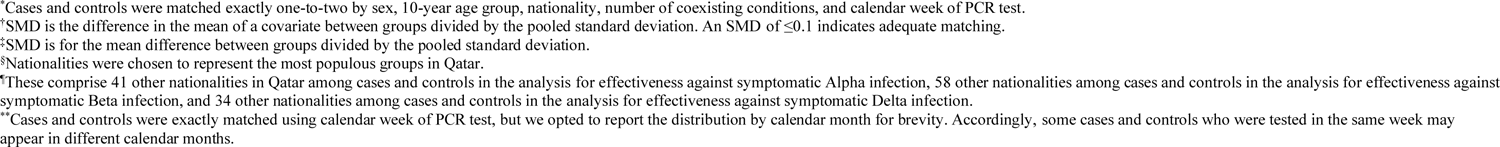
Characteristics of matched cases and controls in samples used to estimate effectiveness against symptomatic Alpha, Beta, or Delta infections in the mRNA-1273 analysis.

**Supplementary Table S3.** Effectiveness of previous pre-Omicron infection, vaccination with mRNA-1273, and hybrid immunity of previous infection and vaccination against symptomatic Alpha, Beta, or Delta infections and against severe, critical, or fatal COVID-19 due to infection with these variants.

| Analyses | Cases<br>(PCR-positive)* |  | Controls<br>(PCR-negative)* |  | Effectiveness against<br>symptomatic infection<br>(95% CI) | Cases<br>(Severe, critical, or fatal<br>COVID-19)† |  | Controls<br>(PCR- negative)‡ |  | Effectiveness against<br>severe, critical, or fatal<br>COVID-19 (95% CI) |
| --- | --- | --- | --- | --- | --- | --- | --- | --- | --- | --- |
|  | Exposed | Unexposed‡ | Exposed | Unexposed‡ |  | Exposed | Unexposed‡ | Exposed | Unexposed‡ |  |
| Alpha symptomatic infection§ |  |  |  |  |  |  |  |  |  |  |
| Previous infection and<br>no vaccination | 28 | 4,562 | 428 | 8,257 | 88.3 (82.8 to 92.1) | 0 | 293 | 63 | 1,010 | 100.0 (94.0 to 100.0)¶ |
| Two doses and no<br>previous infection | 0 | 4,562 | 0 | 8,257 | - | 0 | 293 | 0 | 1,010 | - |
| Two doses and<br>previous infection | 0 | 4,562 | 0 | 8,257 | - | 0 | 293 | 0 | 1,010 | - |
| Three doses and no<br>previous infection | 0 | 4,562 | 0 | 8,257 | - | 0 | 293 | 0 | 1,010 | - |
| Three doses and<br>previous infection | 0 | 4,562 | 0 | 8,257 | - | 0 | 293 | 0 | 1,010 | - |
| Beta symptomatic infection§ |  |  |  |  |  |  |  |  |  |  |
| Previous infection and<br>no vaccination | 97 | 13,459 | 1,340 | 24,096 | 87.5 (84.6 to 89.9) | 1 | 1,133 | 269 | 4,114 | 98.8 (91.4 to 99.8) |
| Two doses and no<br>previous infection | 8 | 13,459 | 276 | 24,096 | 96.0 (91.6 to 98.1) | 0 | 1,133 | 49 | 4,114 | 100.0 (92.2 to 100.0)¶ |
| Two doses and<br>previous infection | 0 | 13,459 | 21 | 24,096 | 100.0 (80.8 to 100.0)¶ | 0 | 1,133 | 2 | 4,114 | 100.0 (-81.2 to 100.0)¶ |
| Three doses and no<br>previous infection | 0 | 13,459 | 0 | 24,096 | - | 0 | 1,133 | 0 | 4,114 | - |
| Three doses and<br>previous infection | 0 | 13,459 | 0 | 24,096 | - | 0 | 1,133 | 0 | 4,114 | - |
| Delta symptomatic infection§ |  |  |  |  |  |  |  |  |  |  |
| Previous infection and<br>no vaccination | 44 | 3,631 | 613 | 5,586 | 90.4 (86.8 to 93.0) | 0 | 125 | 35 | 244 | 100.0 (88.9 to 100.0)¶ |
| Two doses and no<br>previous infection | 477 | 3,631 | 1,521 | 5,586 | 71.0 (66.4 to 74.9) | 3 | 125 | 176 | 244 | 97.9 (91.2 to 99.5) |
| Two doses and<br>previous infection | 11 | 3,631 | 249 | 5,586 | 96.6 (93.5 to 98.2) | 0 | 125 | 19 | 244 | 100.0 (78.6 to 100.0)¶ |
| Three doses and no<br>previous infection | 0 | 3,631 | 15 | 5,586 | 100.0 (72.1 to 100.0)¶ | 0 | 125 | 1 | 244 | 100.0 (-97.4 to 100.0)¶ |
| Three doses and<br>previous infection | 0 | 3,631 | 5 | 5,586 | 100.0 (-8.4 to 100.0)¶ | 0 | 125 | 1 | 244 | 100.0 (-97.4 to 100.0)¶ |

CI denotes confidence interval, COVID-19 coronavirus disease 2019, and PCR polymerase chain reaction.
\*Cases and controls were exactly matched one-to-two by sex, 10-year age group, nationality, number of coexisting conditions, and calendar week of PCR test.
†Cases and controls were exactly matched one-to-five by sex, 10-year age group, nationality, number of coexisting conditions, and calendar week of PCR test.
‡Unexposed was defined as no previous infection and no vaccination.
§A symptomatic infection was defined as a PCR-positive nasopharyngeal swab that was obtained because of the presence of symptoms consistent with a respiratory tract infection. Effectiveness was estimated with the use of a test-negative, case-control study design. COVID-19 severity, criticality, and fatality were defined according to World Health Organization guidelines.
¶The confidence interval was estimated with the use of McNemar's test for matched pairs because of zero events among exposed cases.

